# Clinical characteristics and outcomes of pulmonary haemorrhage in leptospirosis: A retrospective cohort study from Sri Lanka

**DOI:** 10.1101/2024.03.25.24304826

**Authors:** Pramith Ruwanpathirana, Nilanka Perera, Roshan Rambukwella, Dilshan Priyankara

**Affiliations:** Professorial unit in medicine, National Hospital of Sri Lanka, Colombo, Sri Lanka; Department of Medicine, Faculty of Medical Sciences, University of Sri Jayewardenepura, Nugegoda, Sri Lanka; Provincial Director of Health Services, Central Province, Kandy, Sri Lanka; Medical intensive care unit, National Hospital of Sri Lanka, Colombo, Sri Lanka

**Author notes:** Corresponding author –. **Authors’ contributions** DP conceptualized, designed, and conducted the study. RH and PR analysed the data. PR drafted the initial manuscript. NP, DP, and RH critically revised the manuscript. All authors have contributed significantly to the study.

**Keywords:** Leptospirosis, Leptospirosis pulmonary haemorrhage syndrome, plasmapheresis, desmopressin, treatment

## Abstract

**Background:** Leptospirosis, a tropical, spirochaetal infection presents as an acute febrile illness with organ injury. There is a paucity of data on clinical characteristics and treatment for Leptospirosis Pulmonary Haemorrhage Syndrome (LPHS).

**Methodology and principal findings:** A retrospective cohort study was conducted including all patients with LPHS treated in the medical Intensive Care Unit (ICU), at National Hospital Sri Lanka from 2019 to 2022 to describe the clinical characteristics and factors related to poor outcomes. Survival of patients who received different treatment modalities for LPHS was compared.

Seventy patients were studied with a mean age of 42.69 ± 27.84 years and 61 (87.1%) males. Forty-nine (70%) were mechanically ventilated and 16 (22.9%) died. Higher heart rate, higher lactate, lower pH on admission to ICU, and requirement of blood product transfusion were associated with increased mortality. Patients were treated with plasmapheresis (PLEX), intranasal desmopressin (DDAVP), and intravenous steroids alone or in combination as the specific treatment for LPHS. Seven (10%) patients were treated with PLEX alone, 13 (18.6%) with PLEX + DDAVP, 46 (65.7%) with PLEX + DDAVP + steroids, and 4(5.7%) were treated with steroids alone. All patients who received the PLEX and DDAVP combination survived. There were 11 (23.9%) deaths in the PLEX+ DDAVP + steroids group, 3 (49.2%) in the PLEX alone group, and 2 (50%) in the steroids alone group. Mortality was least when PLEX was done for 3 days. Twenty-five (35.7%) patients developed hospital-acquired infections and risk factors were mechanical ventilation and longer ICU stay.

**Conclusions:** The presence of tachycardia, acidosis, and high lactate predicted death in LPHS. PLEX and DDAVP combination had better survival than other treatments alone or in combination for LPHS. Randomized clinical trials are urgently needed to identify the role of PLEX and DDAVP in treating LPHS.

**Author Summary:** Leptospirosis, a tropical infection causes multiple organ injury. Bleeding into the lungs (pulmonary haemorrhage) in leptospirosis is poorly studied. There is no established treatment for pulmonary haemorrhage in leptospirosis (LPHS). A large cohort of LPHS patients treated in a medical ICU in Sri Lanka was studied to identify clinical characteristics and outcomes. The presence of higher heart rate, high lactate, lower pH, need for blood product, and intravenous tranexamic acid were identified as risk factors for death. We compared different treatment modalities used for the treatment of LPHS. In addition to standard treatment with antibiotics and supportive care, a combination of steroids, plasma exchange (PLEX), and intranasal desmopressin (DDAVP) were used to treat LPHS. Mortality was least when patients were treated with DDAVP + PLEX. The optimum duration of PLEX was 3 days. Clinical trials are urgently needed to identify the benefit of PLEX and DDAVP in the treatment of LPHS.

## Introduction

Leptospirosis, a zoonotic spirochaetal infection caused by *Leptospira interrogans*, results in 58,900 deaths (95% CI 23,800–95,900) per annum, worldwide [1,2]. Presenting as an acute fever, leptospirosis can have multi-organ involvement including, acute kidney injury, acute liver injury, myocarditis, aseptic meningitis, and pulmonary haemorrhage. Leptospirosis is classically described as a bi-phasic illness, although it is not always obvious. The initial ‘leptospiraemic phase’ lasts for three to nine days and manifests as a febrile illness with severe myalgia, prostration, and conjunctival suffusion. In this phase, the spirochetes disseminate to the organs haematologically. This is followed by the ‘immune phase’, where IgM against *Leptospira* appears and the organisms disappear from the blood [1]. This phase is characterized by an immune response against the spirochetes residing in the organs such as the liver, lung, kidney, heart, and brain. It is believed that the resulting inflammation leads to the end-organ injury of the immune phase of leptospirosis[3].

Leptospirosis pulmonary haemorrhage syndrome (LPHS) is a poorly studied entity and it is believed to occur in 20% – 70% [4] of patients with a mortality rate of 43%[5] to 55%[6], occurring within the first week of illness[7,8]. Its pathogenesis is postulated to be due to the direct alveolar damage [9] caused by the organism and/or host response against it [10]. Autopsy studies of lungs in LPHS have detected *Leptospira* antigens in the pulmonary vascular endothelium [11, 12], linear deposition of immunoglobulins and complements in alveolar surface [12], foci of interstitial inflammation with lymphocytes, plasma cells, and occasional eosinophils [13]. Studies suggest the occurrence of a cytokine storm in severe leptospirosis [14]. Pro-inflammatory mediators (IL-6, IL-8, and tumour necrosis factor-alpha (TNF∝)) [3] and elevated inducible nitric oxide activity in the lungs were found to be higher in patients who developed LPHS, indicating an immune cell activation [15]. The exact pathogenesis of LPHS is still debatable. Both direct alveolar-capillary damage by the spirochete and immune-mediated damage are equally plausible. In the latter instance, the *Leptospira* antigens found in the lung specimens could be those found in the antigen-antibody complexes.

Treatment of LPHS is not well established due to paucity of evidence. Randomized controlled trials are minimal and most treatment strategies are based on case reports, observational studies, or expert opinion. Steroids and plasma exchange (PLEX) have been tried as treatment modalities considering the immune-based pathogenesis. Many case reports have demonstrated improvement of pulmonary manifestations following steroid treatment in patients with LPHS although a randomized trial conducted in Thailand did not demonstrate a benefit compared to standard of care [16]. The use of steroids was associated with nosocomial infections. PLEX has been shown to significantly improve survival in observational studies with the additional advantage of removing systemic toxic mediators. However, there are no randomized trials supporting this observation. Desmopressin (DDAVP), a selective vasopressin V_2_ agonist has been used in LPHS [17] based on its utility in treating bleeding diathesis in haemophilia A and von Willebrand disease [18].

Sri Lanka is endemic for leptospirosis, reporting many outbreaks with morbidity and mortality. There is an urgent need to generate evidence in an attempt to improve mortality. High-dose steroid therapy (intravenous methylprednisolone followed by oral prednisolone) is recommended in the national guideline of Sri Lanka based on the available limited evidence [19]. PLEX is widely used to treat LPHS in Sri Lanka due to its observed benefit.

We report an observational study on a large number of LPHS patients treated in the Medical Intensive Care Unit (MICU) of the National Hospital of Sri Lanka (NHSL). This is the first report comparing the outcome of multiple modalities of treatment such as steroids, PLEX, DDAVP, or their combinations. This study provides useful information to guide treatment and plan future clinical trials.

## Methods

### Study design and study participants

This retrospective cohort study was done to describe the clinical features, treatment, and disease outcomes of patients with leptospirosis who developed pulmonary haemorrhage. The study was carried out in the MICU, NHSL. All patients admitted to the study site from 1^st^ January 2019 to 1^st^ January 2022 with a microbiological or serological diagnosis of leptospirosis and LPHS were recruited to the study.

### Ethical approval

Ethical approval was obtained from the Ethics Review Committee of the National Hospital of Sri Lanka. Consent was not obtained from participants as this was a retrospective study. Administrative approval from the Deputy Director of the National Hospital of Sri Lanka was obtained to collect anonymized patient details for publication in scientific journals.

### Study procedure

The clinical details were retrospectively extracted from the medical records into a pre-formed data extraction sheet. The socio-demographic details, pre-ICU admission clinical parameters, clinical progression, and treatment given in the ICU were extracted from the medical records.

The definitions used to diagnose leptospirosis and its complications are given in Table 1. The therapeutic decisions were completely under the treating physician’s discretion. Patients were followed up until discharge from the MICU to describe the outcome.

**Table 1:** Definitions used in the study population.

| Condition | Definition |
| --- | --- |
| Microbiologically diagnosed leptospirosis | A positive polymerase chain reaction (PCR) for *Leptospira interrogans* DNA in the serum of the patient within the first 7 days of illness |
| Serologically diagnosed leptospirosis[19] | A titer of $\geq 1:320$ (cut-off recommended for Sri Lanka by the reference laboratory),<br>OR<br>a four-fold rise in titre or seroconversion from acute and convalescent sera<br>using a microscopic agglutination test |
| Pulmonary haemorrhage | Development of haemoptysis (or blood-stained secretions from the endotracheal tube)<br>AND<br>Hypoxia ($SpO_2 < 94\%$)<br>AND<br>Bilateral fluffy opacities in the lung parenchyma in chest radiograph<br>OR<br>HRCT evidence of pulmonary haemorrhage<br>AND<br>Reduction of haemoglobin of more than 1g/dL within 48 hours |
| Myocarditis | Echocardiographic evidence of global hypokinesia<br>AND<br>ECG changes including ST segment and T wave changes not confined to a vascular territory<br>AND<br>Troponin I level higher than 3 times the upper limit of normal (ULN) in the absence of renal impairment and 5 times the ULN in patients with renal impairment |
| Acute kidney injury [20] | • Increase in serum creatinine by 0.3mg/dL or more within 48 hours or<br>• Increase in serum creatinine to 1.5 times baseline or more within the last 7 days or<br>• Urine output less than 0.5 mL/kg/h for 6 hours |

Abbreviations: SpO<sub>2</sub>; peripheral oxygen saturation, HRCT; high resolution computed tomography, ECG; electrocardiograph.

### Statistical analysis

Socio-demographic variables, clinical parameters, complications, treatment, and outcomes were described using descriptive statistics. Percentages were used to describe categorical data. Continuous variables were described using means with standard deviations (SD), and medians with interquartile ranges (IQRs) for variables with a normal or skewed distribution respectively. Differences between categorical variables were assessed using the Chi-square statistic or Fisher’s exact test. Continuous variables with a normal distribution were compared using student t-test or one-way ANOVA. Continuous variables with a skewed distribution were compared using the Mann-Whitney U test or Kruskal-Wallis’s test. Clinical, and laboratory findings and therapy were compared between patients who died and patients who made a complete recovery. The development of hospital-acquired infection and the mortality of the patients were compared between the groups who received different treatment modalities by logistic and Cox regression analysis.

## Results

A total of seventy patients with LPHS were managed in the MICU during the study period. The mean age was 42.69 (±13.92) years with 61 (87.1%) males. Leptospirosis diagnosis was confirmed by polymerase chain reaction in 24 (34.3%) and microscopic agglutination titer in 46 (65.7%). Mechanical ventilation was required for 49 (70%) patients and renal replacement therapy (RRT) was performed in 43 (61.4%) patients with acute kidney injury. There were 16 (22.9%) deaths. Patients who succumbed to the illness had a higher heart rate, lower pH with a lower base excess, and a higher serum lactate on admission to MICU than patients who survived (Table 1). Patients who did not survive the illness, required mechanical ventilation, blood product transfusion, and intravenous tranexamic acid than patients who survived. Mortality was higher in patients who received steroids alone or in combination with other treatments compared to patients who did not receive steroids (Table 2).

**Table 2:**
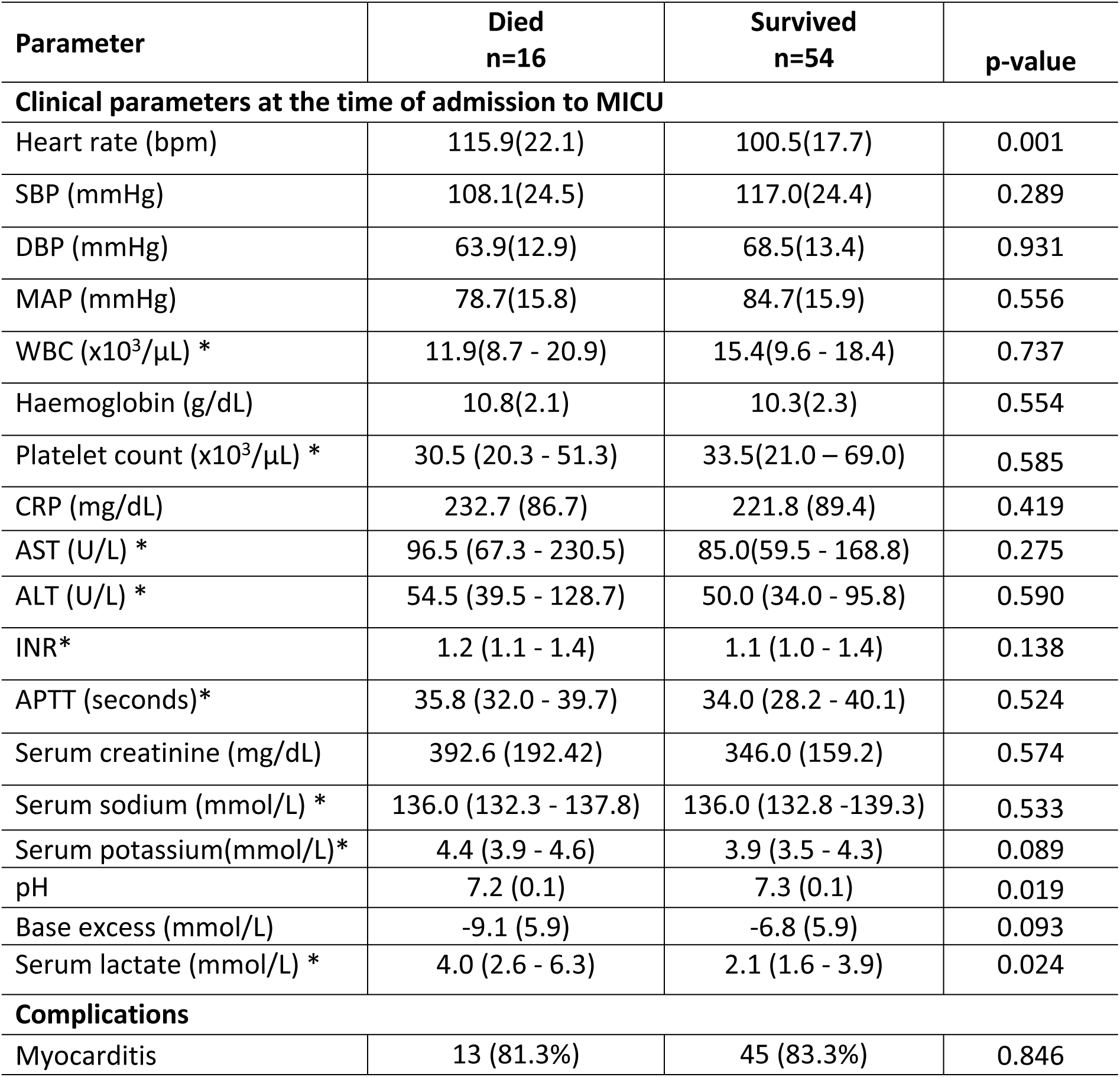
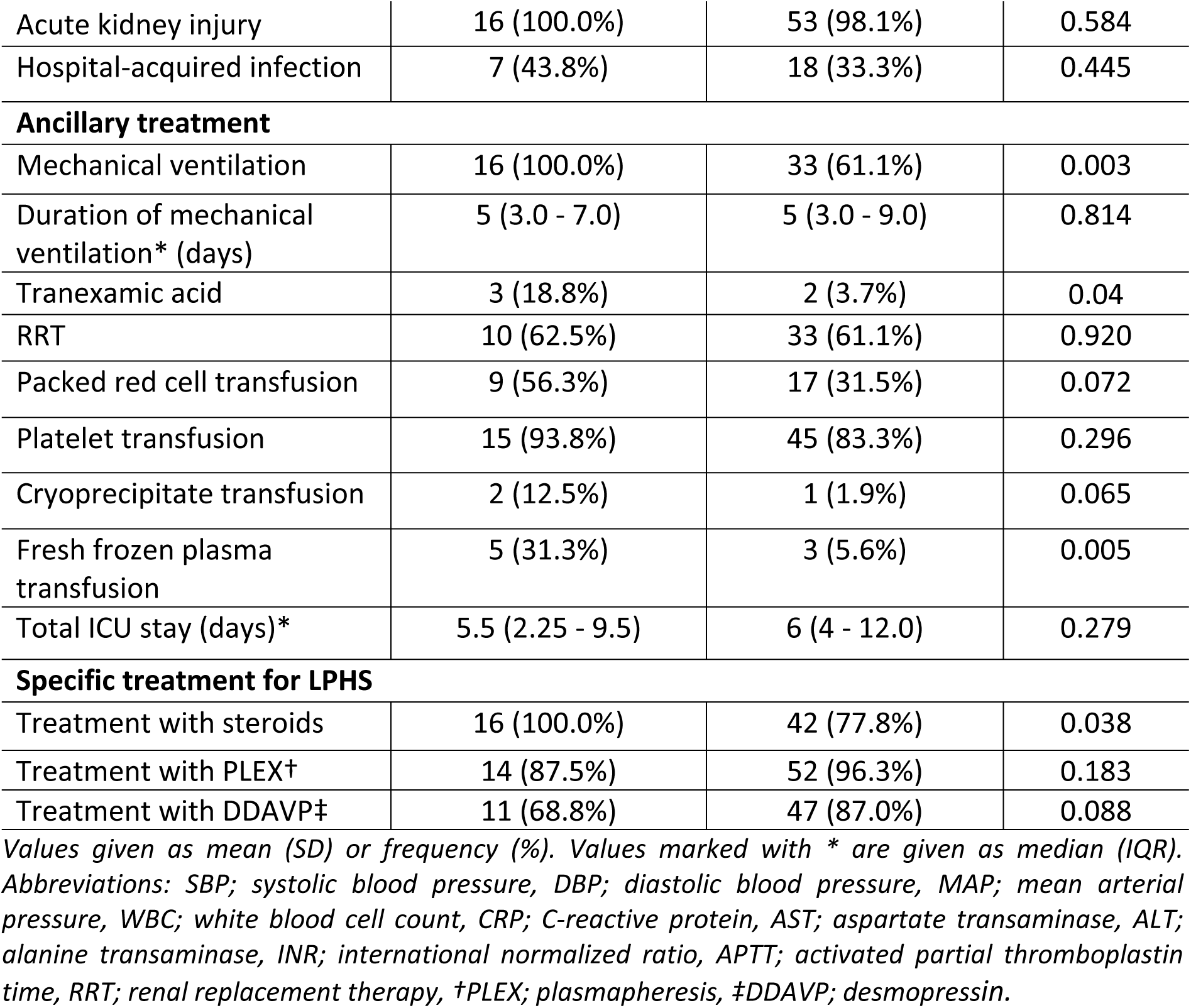
Comparison of clinical and laboratory parameters among patients who died and patients who survived in the study population.

### Treatment modalities for LPHS

Specific treatments received for LPHS were intravenous steroids, intranasal DDAVP (20µg, 6 hourly), and PLEX (60 mL/kg, using fresh frozen plasma, every other day), alone or in combination. The steroids were given as intravenous methylprednisolone (500mg/day - 1g/day) followed by oral prednisolone. The different steroid dosing regimens are given in supplementary table 1 (S1). The characteristics of the patients in the different treatment groups are compared in Table 3. Patient characteristics and laboratory findings revealed that the group who received PLEX alone had a significantly higher heart rate, lower systolic blood pressure, higher serum lactate levels, and higher serum creatinine than the other groups. In addition, a higher proportion in this group received fresh frozen plasma, cryoprecipitate, and packed red cell transfusions. Other laboratory parameters were not significantly different in the treatment groups.

**Table 3:** Comparison of patient characteristics, treatment, and outcomes between treatment strategies for LPHS.

| Parameter | Total<br>(n=70) | Treatment modalities |  |  |  |  |
| --- | --- | --- | --- | --- | --- | --- |
|  |  | PLEX†<br>alone<br>(n=7) | PLEX† +<br>DDAVP‡<br>(n=13) | PLEX† +<br>DDAVP‡ +<br>Steroids<br>(n=46) | Steroids<br>alone<br>(n=4) | p-value |
| Clinical parameters at the time of admission to MICU |  |  |  |  |  |  |
| Heart rate (bpm) | 104.0 (19.7) | 117.3 (28.5) | 87.2 (14.5) | 106.4(17.4) | 107.5 (14.4) | 0.002 |
| SBP (mmHg) | 115.0 (24.5) | 108.3 (22.6) | 122.5<br>(21.0) | 114.1<br>(26.5) | 112.5<br>(12.6) | 0.616 |
| DBP (mmHg) | 67.4<br>(13.3) | 63.1<br>(7.4) | 68.7<br>(9.1) | 67.1<br>(15.1) | 75.0<br>(9.1) | 0.546 |
| MAP (mmHg) | 83.3<br>(16.0) | 78.2<br>(12.2) | 86.6<br>(12.6) | 82.8<br>(17.8) | 87.5<br>(8.8) | 0.666 |
| WBC (x10<sup>3</sup>/μL) * | 14.2<br>(9.5 - 18.4) | 12.6<br>(11.8 - 17.1) | 15.5<br>(11.0 - 23.3) | 12.7 (7.8 - 9.2) | 14.1 (10.0 - 25.3) | 0.839 |
| Haemoglobin (g/dL) | 10.4 (2.2) | 10.1 (2.7) | 10.8 (1.5) | 10.4 (2.4) | 10.6 (1.3) | 0.884 |
| Platelet count<br>(x10<sup>3</sup>/μL) * | 32.0<br>(21.0 - 64.3) | 51.0<br>(34.0 – 60.0) | 23.0<br>(20.0 - 82.5) | 32.0 (23.8 - 65.8) | 19.0 (16.3 - 38.3) | 0.378 |
| CRP (mg/dL) | 224.3<br>(88.3) | 177.6<br>(48.8) | 203.3<br>(75.9) | 234.3<br>(92.5) | 260.5<br>(112.5) | 0.269 |
| AST (U/L) * | 89.5<br>(61.0 – 178.0) | 101.0<br>(63 – 166) | 79.0 (44.5 - 225.5) | 90.5 (65.5 – 178) | 76.5 (62.5 - 205.25) | 0.881 |
| ALT (U/L) * | 51.0 (36.25 - 95.75) | 45.0<br>(34.0 – 134.0) | 45.0 (33.5 – 100.0) | 53.0 (36.3 - 90.5) | 49.5 (41.0 – 94.0) | 0.964 |
| INR* | 1.1<br>(1.0 - 4.4) | 1.0<br>(0.9 - 1.4) | 1.1<br>(1.1 -1.4) | 1.1<br>(1.0 -1.4) | 1.3<br>(1.1 - 1.9) | 0.485 |
| APTT (seconds)* | 34.4<br>(29.6 - 40.1) | 33.0<br>(26.0 – 35.0) | 38.0<br>(29.3 -42.3) | 34.9 (29.6 - 40.1) | 31.1<br>(29.3 - 41.9) | 0.503 |
| Serum creatinine<br>(μmol/L) | 356.7 (167.1) | 428.6 (191.1) | 331.8<br>(178.7) | 361.1<br>(162.9) | 261.3<br>(126.2) | 0.411 |
| Serum sodium<br>(mmol/L) * | 136.0 (132.7-139) | 135.0<br>(131.0 – 136.0) | 136.0 (128-140.5) | 136.0<br>(133.0 – 139.0) | 137.5 (133.3 - 140.3) | 0.663 |
| Serum potassium(mmol/L) * | 4.0<br>(3.5 - 4.4) | 4.3<br>(3.2 - 4.6) | 3.6<br>(3.2 - 3.95) | 4.0<br>(3.6 - 4.5) | 4.3<br>(3.80 - 5.15) | 0.122 |
| pH | 7.3 (0.1) | 7.2 (0.2) | 7.4 (0.1) | 7.3 (0.1) | 7.2 (0.2) | 0.010 |
| Base excess (mmol/L) | -7.3 (6.0) | -5.3 (5.7) | -5.5 (6.2) | -7.8 (6.0) | -10.7 (4.7) | 0.323 |
| Serum lactate (mmol/L) * | 2.6<br>(1.6 - 4.3) | 3.8<br>(1.4 - 5.3) | 2.1<br>(1.15 -2.95) | 2.6<br>(1.7 - 4.8) | 2.6<br>(1.325 - 6.8) | 0.502 |
| **Complications** |  |  |  |  |  |  |
| Myocarditis | 58<br>(82.9%) | 4<br>(57.1%) | 12<br>(92.3%) | 39<br>(84.8%) | 3<br>(75.0%) | 0.224 |
| Acute kidney injury | 69<br>(98.6%) | 7<br>(100.0%) | 13<br>(100.0%) | 45<br>(97.8%) | 4<br>(100.0%) | 0.912 |
| **Ancillary treatment** |  |  |  |  |  |  |
| Mechanical ventilation | 49<br>(70.0%) | 5<br>(71.4%) | 6<br>(46.2%) | 35<br>(76.1%) | 3<br>(75.0%) | 0.223 |
| Tranexamic acid | 5<br>(7.1%) | 1<br>(14.3%) | 0<br>(0.0%) | 3<br>(6.5%) | 1<br>(25.0%) | 0.322 |
| Renal replacement therapy | 43<br>(61.4%) | 4<br>(57.14%) | 6<br>(46.2%) | 32<br>(69.6%) | 1<br>(25.0%) | 0.182 |
| **Blood product transfusion** |  |  |  |  |  |  |
| Packed red cell transfusion | 26<br>(37.1%) | 4<br>(57.1%) | 0<br>(0.0%) | 21<br>(45.7%) | 1<br>(25.0%) | 0.014 |
| Platelet transfusion | 60<br>(85.7%) | 6<br>(85.7%) | 9<br>(69.2%) | 43<br>(93.5%) | 2<br>(50.0%) | 0.025 |
| Cryoprecipitate transfusion | 3<br>(4.3%) | 1<br>(14.3%) | 0<br>(0.0%) | 2<br>(4.4%) | 0<br>(0.0%) | 0.481 |
| Fresh frozen plasma transfusion | 8<br>(11.4%) | 3<br>(42.9%) | 0<br>(0.0%) | 4<br>(8.7%) | 1<br>(25.0%) | 0.023 |
| **Outcome** |  |  |  |  |  |  |
| Death at 32 days | 16<br>(22.9%) | 3<br>(42.9%) | 0<br>(0.0%) | 11<br>(23.9%) | 2<br>(50.0%) | 0.068 |
| Duration of mechanical ventilation*/days | 5<br>(3.0 - 8.0) | 3<br>(2.0 - 8.5) | 3<br>(2.5 - 4.5) | 5<br>(4.0 - 8.5) | 1<br>(1) | 0.194 |
| Duration of treatment in the ICU*/days | 6<br>(3.0 - 11.0) | 5<br>(3.0 - 6.0) | 5<br>(3.0 - 10.5) | 6<br>(4.0- 11.25) | 2<br>(1.0 - 22.5) | 0.249 |
| Hospital-acquired infection | 25<br>(35.7%) | 2<br>(28.6%) | 1<br>(7.7%) | 21<br>(45.7%) | 1<br>(25.0%) | 0.079 |

Mean (SD) or frequency (%) used. Nonparametric distributions are indicated with \* and given as median (IQR). Abbreviations: PCR; Polymerase chain reaction, MAT; microscopic agglutination test, SBP; systolic blood pressure, DBP; diastolic blood pressure, MAP; mean arterial pressure, WBC; white blood cell count, CRP; C reactive protein, AST; aspartate transaminase, ALT; alanine transaminase, INR;
international normalized ration, APTT; activated partial thromboplastin time, RRT; renal replacement therapy, †PLEX; plasmapheresis, ‡DDAVP; desmopressin

All patients treated with the PLEX and DDAVP combination survived. Mortality was low in the group treated with PLEX + DDAVP + steroids compared to PLEX alone or steroids alone. A logistic regression and a univariate Cox regression analysis were performed to calculate the odds and hazard ratios for death among different treatment modalities (Figure 1a) and treatment combinations (Figure 1b) using the group treated with steroids as the reference (Table 4). The odds ratio (OR) for the group that received PLEX + DDAVP without steroids could not be calculated as the morality was 0%.

**Figure 1:**
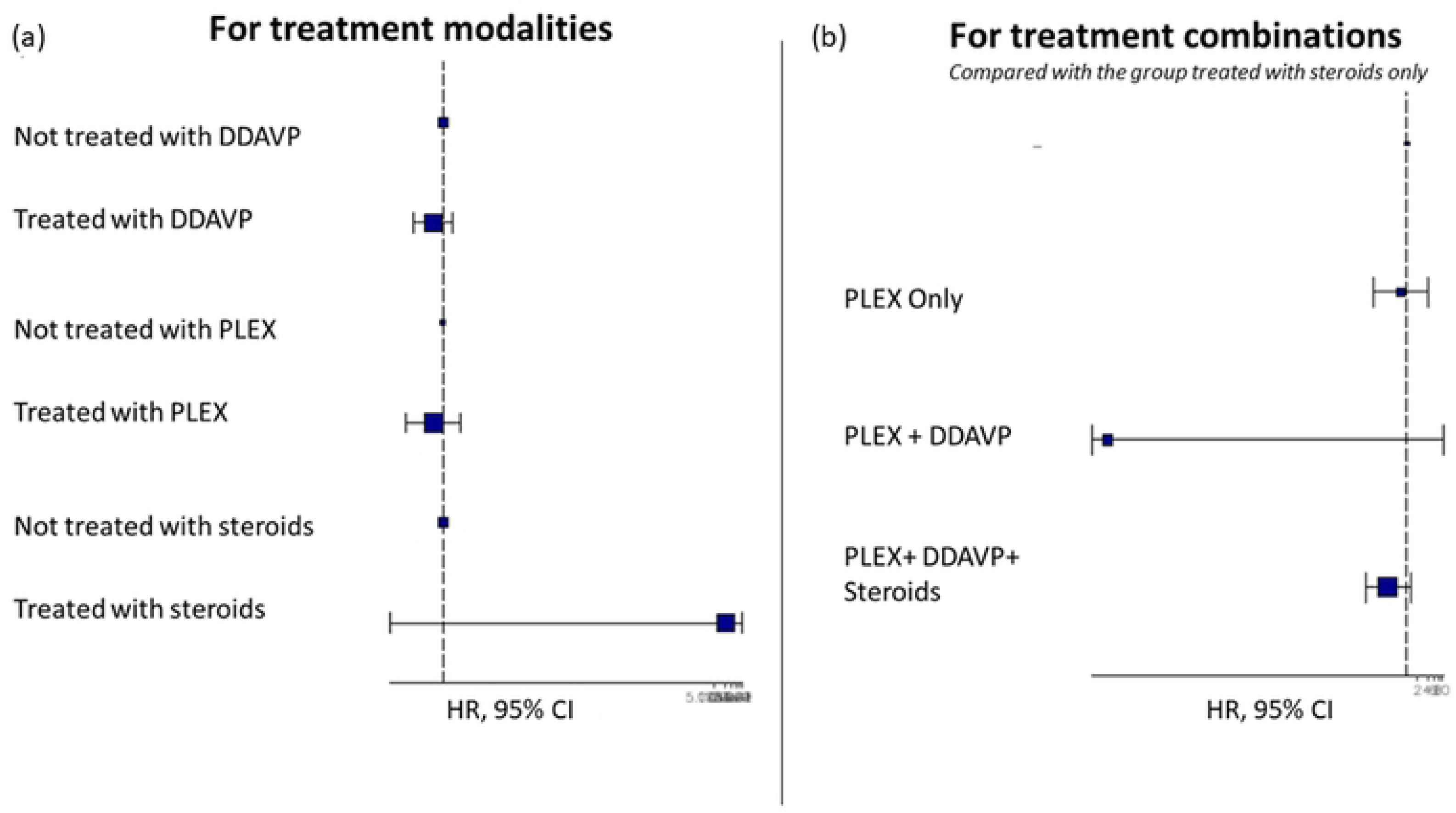
Hazard regression plots for mortality in patients with LPHS. Cox regression analysis was performed to analyze the effect of different treatments received by patients with LPHS on mortality. Patients were treated with plasmapheresis (PLEX), desmopressin (DDAVP), and steroids alone or in different combinations. Hazard regression plots demonstrate the risk of mortality (hazard ratio) and the confidence interval. (a) Patients who received each treatment modality were compared with patients who did not receive the treatment. (b) Treatment combinations were compared with the group who received steroids alone.

**Table 4:** Risk of *mortality* in patients with LPHS who received different treatment combinations.

| Treatment | Total<br>n=70 | Mortality<br>n (%) | OR (95% CI) | HR (95% CI) |
| --- | --- | --- | --- | --- |
| PLEX alone | 7 | 3 (42.9%) | 0.75 (0.06 – 8.83) | 0.65 (0.11 – 3.92) |
| PLEX + DDAVP | 13 | 0 (0%) | NC | NC |
| PLEX + DDAVP + steroids | 46 | 11 (23.9%) | 0.31 (0.04 – 2.5) | 0.29 (0.06 – 1.32) |
| Steroids alone | 4 | 2 (50%) |  |  |

Abbreviations: OR; odds ratio, HR; Hazards ratio, NC; Not calculated

Transfusion of blood products and treatment with tranexamic acid did not meet statistical significance in the logistic regression analysis predictors of mortality. The Kaplan-Meier survival curves comparing the treatment combinations in patients with LPHS are given in Figure 2. All patients who were discharged to the ward after complete recovery were assumed to have survived till the end of follow up i.e.: 32 days (the maximum duration of ICU stay before death).

**Figure 2:**
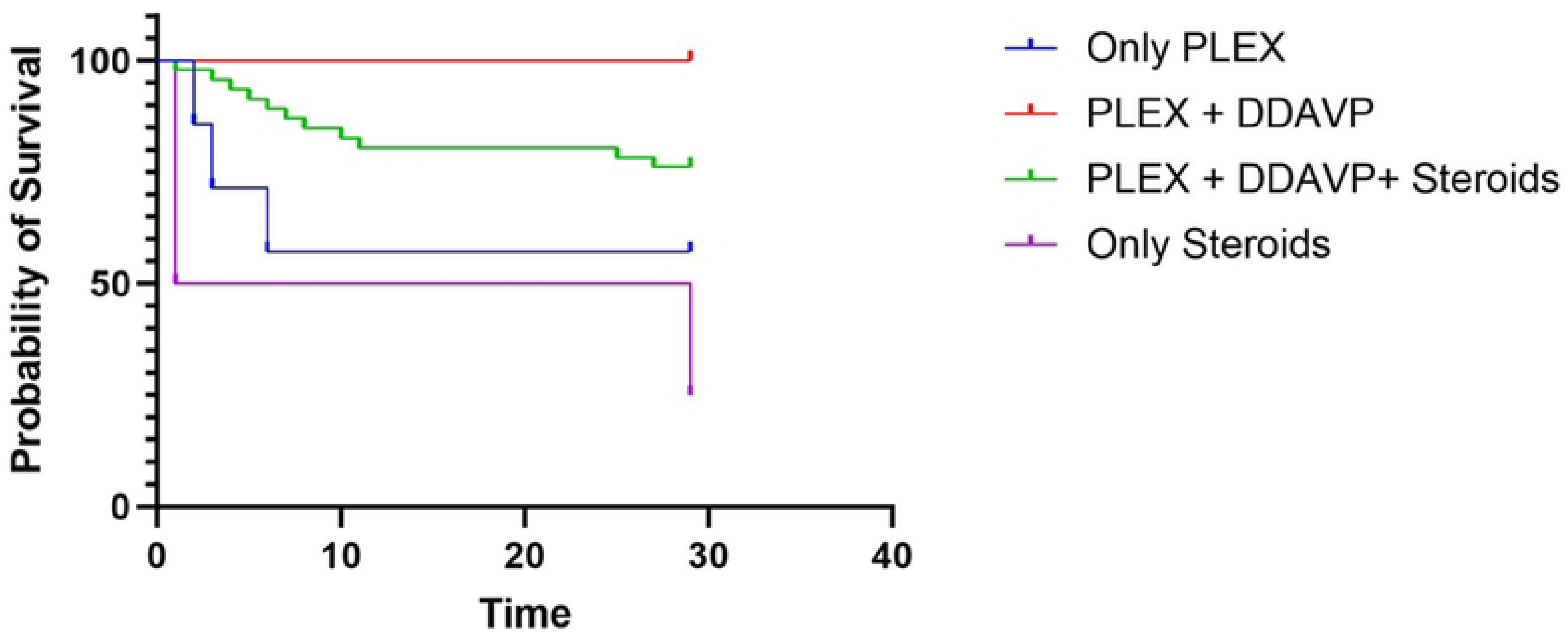
Kaplan-Meier survival curves of patients with LPHS who received different treatment combinations. The figure shows the survival of patients with LPHS who received plasmapheresis (PLEX) alone (n=7), PLEX and desmopressin (DDAVP) combination (n=13), PLEX+DDAVP and steroids combination (n=46) or steroids alone (n=4).

A Cox regression analysis was performed to assess the relationship of the duration of PLEX with mortality. Since PLEX for 1 day and 6 days were done only for 1 patient each, and PLEX for 2 days had only 2 patients, they were excluded from the analysis. The hazard ratios for death were calculated for different durations of PLEX compared to the group who received PLEX for three days. Hazard ratios were 2.46 (0.58-10.47, p>0.05) and 5.9 (1.13-30.95, p 0.04) in the groups who received PLEX for 4 days and 5 days respectively. Steroids (p=0.620) and DDAVP (p=0.430) were used equally among the groups who received different durations of PLEX. The Kaplan-Meier survival curves comparing the duration of PLEX are given in Figure 3.

**Figure 3:**
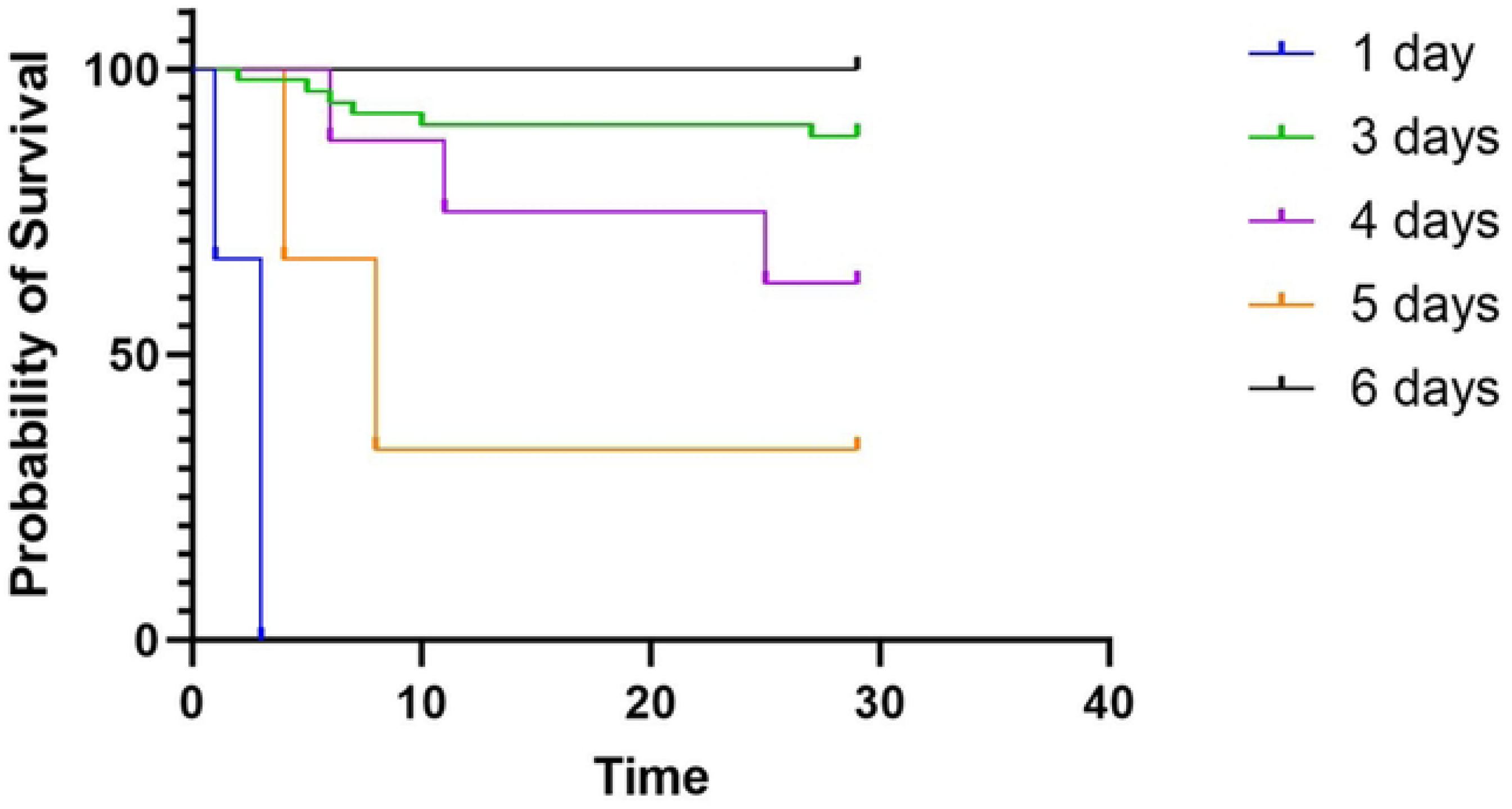
Kaplan-Meier survival curves of patients with LPHS who received different durations of plasmapheresis (PLEX). Figure shows the survival of patients with LPHS who received PLEX for 1 day (n=1), 2 days (n=2), 3 days (n=54), 4 days (n=8), 5 days (n=3) and 6 days (n=1).

### Hospital-acquired infection (HAI) in the study population

HAI occurred in 25 (35.7%) patients. Respiratory tract infections (n=16, 61.5%) and catheter-associated bloodstream infections (n=2, 7.7%) were the common sources of HAIs. Logistic regression analysis was performed to identify risk factors for the development of HAI. Mechanical ventilation (OR 30.34, CI 1.55- 592.10) and duration of ICU stay (OR 1.37, CI 1.12- 1.68) were risk factors for the development of HAI. A higher proportion of patients in the PLEX + DDAVP+ steroid group developed HAI compared to the groups who received PLEX + DDAVP combination or PLEX and steroids alone (Table 3). However, different treatment modalities received by patients for LPHS (compared to receiving steroids alone), renal replacement therapy, or blood product transfusion were not significantly associated with developing HAI in logistic regression analysis.

## Discussion

LPHS is a dreaded complication of leptospirosis due to its high mortality and challenges in management. There is a paucity of data on the clinical characteristics and outcomes of LPHS. In this retrospective cohort study of a large number of LPHS patients, we report characteristics, risk factors for death, and outcomes from different treatment modalities for LPHS. This is the first study that compares the outcome of PLEX, DDAVP, and steroid treatment alone or in combination for LPHS. Findings from this study provide a platform to design future clinical trials.

Patients who developed LPHS requiring ICU care, had myocarditis (n=58, 82.9%), acute kidney injury (n=69, 98.6%), and modest elevation in liver enzymes. A significant proportion required renal replacement therapy (n=43, 61.4%), mechanical ventilation (n=49, 70%), and blood product transfusion (n=31, 44.3%). A previous Sri Lankan study reported a 35.4% mortality in LPHS[21]. A mortality of 22.9% observed in this study is relatively lower than previous reports from Sri Lanka and other countries [22]. We found that demographic factors such as age and gender were not significantly different in the patients who died compared to patients who survived. However, tachycardia, lactataemia, and acidosis on admission to MICU were more common in the patients who died compared to the survivors. A higher proportion of patients who succumbed to the illness were mechanically ventilated and received intravenous tranexamic acid and fresh frozen plasma. This observation suggests that severe pulmonary haemorrhage leading to respiratory failure and ongoing bleeding with coagulopathy would have been present in this group.

Specific treatment for LPHS is controversial. Literature is scarce and any evidence supporting steroids, PLEX, or DDAVP is based on case reports, observational studies, and expert opinion. With the emergence of LPHS cases throughout the country, many centres in Sri Lanka use PLEX as a therapeutic measure. Evidence for PLEX was based on the benefit observed in the treatment of pulmonary haemorrhage in autoimmune diseases such as Goodpasture’s syndrome, ANCA-associated vasculitis, systemic lupus erythematosus (SLE), and other autoimmune reno-pulmonary syndromes [23–25]. The above diseases arise due to either direct deposition of autoantibodies or antigen-antibody complexes in the alveolar-capillary membrane, resulting in inflammation, pulmonary vasculitis, tissue damage, necrosis, and pulmonary haemorrhage. Removal of auto-antibodies and circulating antigen-antibody complexes provide the rationale for PLEX in the above conditions. As immune-mediated damage is one theory for the pathogenesis of LPHS, it can be postulated that PLEX might be an effective therapy. In a previous study of 80 patients with LPHS at Teaching Hospital Karapitiya Sri Lanka, 59 (73.75%) underwent PLEX of which 19 died (fatality rate; 32.2%) compared to 17 (fatality rate; 89.5%) in the group which did not undergo PLEX [7]. Furthermore, the survival rate was higher (*n* = 35, 74.5%) when the PLEX was performed within the first 48 hours compared to patients in whom the procedure was done after 48 hours (*n* = 5, 54.5%). PLEX was done for a mean duration of 3 consecutive days, using fresh frozen plasma. In an Indian study, a survival benefit was noted when two cycles of PLEX 24 hours apart were combined with IV cyclophosphamide, to treat LPH[26]. Numerous case reports have described successful treatment of LPHS with PLEX, alone or in combination with steroids, extra-corporeal membrane oxygenation [15,27–29]. All patients who received PLEX+DDAVP survived. There was no statistically significant difference in the hazard ratio between different treatment combinations of LPHS when the PLEX+DDAVP group was excluded from the analysis. The PLEX + DDAVP group was excluded as its mortality was zero. However, survival curves demonstrated a favourable outcome in patients who received PLEX + DDAVP combination than the other treatment groups. The PLEX alone group had higher lactate and acidosis than the other groups on admission to MICU. As these parameters were predictors of mortality subsequently, the benefit of PLEX could have been masked by initial severe LPHS in the group. The optimal duration of PLEX is either three or four days according to the study findings.

Intravenous bolus doses of corticosteroids are used to treat pulmonary haemorrhages in auto-immune diseases. Using the same argument as for PLEX, corticosteroids have been used to treat LPHS. In the year 2008, Sri Lanka recorded a surge of leptospirosis cases with 7406 hospital admissions, 217 deaths, and a case fatality rate of 3%. Kularatne *et al* treated patients with severe leptospirosis with intravenous methyl-prednisolone 500 mg for 3 days followed by 8 mg orally for 5 days. Treatment was compared to a historical cohort without steroid treatment [30]. The outcome was analysed based on the organ dysfunction, co-morbidities, and steroids significantly (p=0.012) reduced mortality in patients with LPHS. A mortality benefit was noted in the patients who received steroids in another study [31]. However, this effect was seen only if the steroids were given within the first 12 hours. In an Australian study describing the intensive care treatment for severe leptospirosis, IV hydrocortisone 200mg daily was used as a supportive measure [32]. There was no significant difference in the incidence of pulmonary haemorrhage between the patients who did and did not receive steroids. The mortality did not differ between the two groups. However, the patients treated with steroids were sicker with a high APACHE III score and required more vasopressor support. As per PLEX, numerous case reports describe successful management of LPHS with high-dose bolus steroids. A concern of steroid treatment has been the increased incidence of HAIs. In our study, univariate analysis revealed that steroid treatment compared to no steroids, increased mortality. This observation was not statistically significant on Cox regression analysis. However, the addition of steroids to PLEX+DDAVP reduced survival compared to PLEX+DDAVP alone. In addition, there was a non-significant increase of HAIs in steroid-treated groups compared to the PLEX+ DDAVP group.

Evidence for DDAVP in LPHS is limited. DDAVP releases endothelial haemostatic factors thereby, shortens prolonged bleeding times, and enhancing platelet adhesiveness to injured vessels[18] by activation and secretion of von Willebrand (vWF) factor from the endothelial cells. Leptospirosis is associated with bleeding beyond the lung ranging from petechiae, ecchymoses, and epistaxis to massive gastro-intestinal bleeds. A platelet dysfunction is known to occur in leptospirosis [33]. Furthermore, a coagulopathy associated with leptospirosis has been described. In an animal study, *Leptospira* infected hamsters had a prolonged PT, APTT, and a thrombin time in all organ extracts, especially in the lung and the liver, probably due to interference with the prothrombinase complex [34]. Platelet dysfunction, coagulopathy, and thrombocytopenia might be contributing to the increased bleeding observed in leptospirosis. DDAVP might be acting by increasing the platelet activation and local production and secretion of vWF and factor VIII. A theoretical risk of causing, aggravating disseminated intravascular coagulation is present with the use of DDAVP in severe leptospirosis. Pea *et al* described six patients with LPHS treated with intravenous DDAVP [35]. Bleeding from the endotracheal tube had reduced in intubated patients within one hour of DDAVP infusion, despite repeated aspiration. An open randomized controlled trial of high-dose dexamethasone and intravenous desmopressin alone or in combination did not alter the outcome compared to placebo [16]. Intranasal DDAVP in LPHS has not been studied previously. Further studies are needed to explore its efficacy. In the interim, it can be stated that DDAVP seems to be effective in combination with PLEX as a treatment for LPHS.

There was no statistically significant difference in mortality between different treatment groups when the PLEX+DDAVP group was excluded. However, our results demonstrate a clear survival benefit in PLEX+ DDAVP treatment compared to the addition of steroids, PLEX alone, or steroids alone. Reduced overall mortality in the study population compared to previous literature could also be due to the use of such therapeutic measures. Differences in the severity of the underlying disease could have played a role in interpreting the findings of this study. As such, appropriately designed randomized controlled trials are urgently needed to generate evidence and improve patient outcomes.

Patients who developed HAIs had a higher heart rate, INR, and a lower base excess. They were mechanically ventilated for a longer period, and treated with steroids and renal replacement therapy compared to the group that did not develop HAIs. Transfusion (FFP, RCC, platelet) rates were higher in the group that developed HAIs. However, only mechanical ventilation and duration of ICU stay were predictive of the development of HAI.

Treatment of LPHS is challenging due to the paucity of evidence. We have compared PLEX, DDAVP, and the use of steroids in the management of LPHS alone or in combination. A major limitation of this study is that the treatment was solely under the treating physician’s discretion. Hence, the comparison between the treatment strategies is not ideal. Similarly, the duration of PLEX and the steroid regimens used were not uniform. Furthermore, the extent of lung involvement in LPHS has not been described or compared in this study and poses a limitation in interpreting the results. However, the important observations of this cohort of large numbers of LPHS patients lay a foundation to conduct future studies and randomized controlled trials.

## Conclusions

Patients admitted with LPHS to an intensive care setting had renal, liver, and myocardial involvement requiring organ support, renal replacement therapy, and blood product transfusion. The presence of tachycardia, acidosis, and high lactate predicted death in LPHS. PLEX and DDAVP combination had better survival than other treatments alone or in combination for LPHS. Randomized clinical trials are urgently needed to identify the role of PLEX and DDAVP in treating LPHS.

## Data Availability

Data are available in the BioStudies database (https://www.ebi.ac.uk/biostudies/) under accession number S-BSST1368.

## Abbreviations

AKI: acute kidney injury
DDAVP: desmopressin
FFP: Fresh frozen plasma
HR: hazard ratio
ICU: intensive care unit
LPHS: Leptospirosis pulmonary haemorrhage syndrome
MAT: microscopic agglutination test
MICU: Medical intensive unit
NHSL: National Hospital of Sri Lanka
OR: odds ratio
PCR: polymerase chain reaction
PLEX: plasma exchange
RCC: Red cell concentrate
RRT: renal replacement therapy
ULN: upper limit of normal
vWF: von Willebrand factor

## Acknowledgments

The authors would like to acknowledge the staff of MICU, National Hospital of Sri Lanka for their assistance in conducting the study.

## Supporting information

S1 Table: Different regimens of steroids used to treat LPHS

## Declarations

### Ethics approval and consent to participate

Ethical approval was obtained from the Ethics Review Committee of the National Hospital of Sri Lanka

### Consent for publication

As this was a retrospective study using the patient’s clinical details, consent was not obtained. All collected data were anonymised. Details that would point toward patient identification were not collected. Administrative approval from the Deputy Director of the National Hospital of Sri Lanka was obtained for the collection of anonymised patient details and publication in scientific journals.

## References

1. Rajapakse S. Leptospirosis: clinical aspects. Clinical Medicine. 2022;22: 14. doi:10.7861/CLINMED.2021-0784

2. Costa F, Hagan JE, Calcagno J, Kane M, Torgerson P, Martinez-Silveira MS, et al. Global Morbidity and Mortality of Leptospirosis: A Systematic Review. PLoS Negl Trop Dis. 2015;9. doi:10.1371/JOURNAL.PNTD.0003898

3. Cagliero J, Villanueva SYAM, Matsui M. Leptospirosis pathophysiology: Into the storm of cytokines. Front Cell Infect Microbiol. 2018;8: 274770. doi:10.3389/FCIMB.2018.00204/BIBTEX

4. Carvalho CRR, Bethlem EP. Pulmonary complications of leptospirosis. Clin Chest Med. 2002;23: 469–478. doi:10.1016/S0272-5231(01)00010-7

5. Singh SS, Vijayachari P, Sinha A, Sugunan AP, Rasheed MA, Sehgal SC. Clinico-epidemiological study of hospitalized cases of severe leptospirosis. Indian J Med Res. 1999;109: 94–99.

6. Marotto PCF, Nascimento CMR, Eluf-Neto J, Marotto MS, Andrade L, Sztajnbok J, et al. Acute lung injury in leptospirosis: clinical and laboratory features, outcome, and factors associated with mortality. Clin Infect Dis. 1999;29: 1561–1563. doi:10.1086/313501

7. Herath N, Uluwattage W, Weliwitiya T, Karunanayake L, Lekamwasam S, Ratnatunga N, et al. Sequel and therapeutic modalities of leptospirosis associated severe pulmonary haemorrhagic syndrome (SPHS); A Sri Lankan experience. BMC Infect Dis. 2019;19: 1–8. doi:10.1186/S12879-019-4094-0/TABLES/7

8. Fonseka CL, Dahanayake NJ, Mihiran DJD, Wijesinghe KM, Liyanage LN, Wickramasuriya HS, et al. Pulmonary haemorrhage as a frequent cause of death among patients with severe complicated Leptospirosis in Southern Sri Lanka. PLoS Negl Trop Dis. 2023;17: e0011352. doi:10.1371/JOURNAL.PNTD.0011352

9. Luks AM, Lakshminarayanan S, Hirschmann J V. Leptospirosis presenting as diffuse alveolar hemorrhage: case report and literature review. Chest. 2003;123: 639–643. doi:10.1378/CHEST.123.2.639

10. Dolhnikoff M, Mauad T, Bethlem EP, Carvalho CRR. Pathology and pathophysiology of pulmonary manifestations in leptospirosis. Braz J Infect Dis. 2007;11: 142–148. doi:10.1590/S1413-86702007000100029

11. Nicodemo AC, Duarte MIS, Alves VAF, Takakura CFH, Santos RTM, Nicodemo EL. Lung lesions in human leptospirosis: microscopic, immunohistochemical, and ultrastructural features related to thrombocytopenia. Am J Trop Med Hyg. 1997;56: 181–187. doi:10.4269/AJTMH.1997.56.181

12. Croda J, Neto AND, Brasil RA, Pagliari C, Nicodemo AC, Duarte MIS. Leptospirosis pulmonary haemorrhage syndrome is associated with linear deposition of immunoglobulin and complement on the alveolar surface. Clinical Microbiology and Infection. 2010;16: 593–599. doi:10.1111/J.1469-0691.2009.02916.X

13. Arean VM. The Pathologic Anatomy and Pathogenesis of Fatal Human Leptospirosis (Weil’s Disease). Am J Pathol. 1962;40: 393.

14. Reis EAG, Hagan JE, Ribeiro GS, Teixeira-Carvalho A, Martins-Filho OA, Montgomery RR, et al. Cytokine Response Signatures in Disease Progression and Development of Severe Clinical Outcomes for Leptospirosis. PLoS Negl Trop Dis. 2013;7: e2457. doi:10.1371/JOURNAL.PNTD.0002457

15. Chen HI, Kao SJ, Hsu YH. Pathophysiological mechanism of lung injury in patients with leptospirosis. Pathology. 2007;39: 339–344. doi:10.1080/00313020701329740

16. Niwattayakul K, Kaewtasi S, Chueasuwanchai S, Hoontrakul S, Chareonwat S, Suttinont C, et al. An open randomized controlled trial of desmopressin and pulse dexamethasone as adjunct therapy in patients with pulmonary involvement associated with severe leptospirosis. Clinical Microbiology and Infection. 2010;16: 1207–1212. doi:10.1111/J.1469-0691.2009.03037.X

17. Pea L, Roda L, Boussaud V, Lonjon B. Desmopressin therapy for massive hemoptysis associated with severe leptospirosis. Am J Respir Crit Care Med. 2003;167: 726–728. doi:10.1164/RCCM.200205-450CR

18. Mannucci PM. Desmopressin (DDAVP) in the Treatment of Bleeding Disorders: The First 20 Years. Blood. 1997;90: 2515–2521. doi:10.1182/BLOOD.V90.7.2515

19. National Guidelines on Management of Leptospirosis Epidemiology Unit Ministry of Health, Nutrition and Indigenous Medicine Sri Lanka. 2016.

20. KDIGO Clinical Practice Guideline for Acute Kidney Injury. doi:10.1038/kisup.2012.1

21. Fonseka CL, Dahanayake NJ, Mihiran DJD, Wijesinghe KM, Liyanage LN, Wickramasuriya HS, et al. Pulmonary haemorrhage as a frequent cause of death among patients with severe complicated Leptospirosis in Southern Sri Lanka. PLoS Negl Trop Dis. 2023;17. doi:10.1371/JOURNAL.PNTD.0011352

22. Dolhnikoff M, Mauad T, Bethlem EP, Carvalho CRR. Pathology and pathophysiology of pulmonary manifestations in leptospirosis. Braz J Infect Dis. 2007;11: 142–148. doi:10.1590/S1413-86702007000100029

23. Simpson IJ, Doak PB, Williams LC, Blacklock HA, Hill RS, Teague CA, et al. Plasma exchange in Goodpasture’s syndrome. Am J Nephrol. 1982;2: 301–311. doi:10.1159/000166666

24. Klemmer PJ, Chalermskulrat W, Reif MS, Hogan SL, Henke DC, Falk RJ. Plasmapheresis Therapy for Diffuse Alveolar Hemorrhage in Patients with Small-Vessel Vasculitis. American Journal of Kidney Diseases. 2003;42: 1149–1153. doi:10.1053/j.ajkd.2003.08.015

25. Abe Y, Kusaoi M, Tada K, Yamaji K, Tamura N. Efficacy of plasma exchange therapy for diffuse alveolar hemorrhage in patients with microscopic polyangiitis. Therapeutic Apheresis and Dialysis. 2022;26: 515–521. doi:10.1111/1744-9987.13824

26. Trivedi S V, Vasava AH, Bhatia LC, Patel TC, Patel NK, Patel NT. Plasma exchange with immunosuppression in pulmonary alveolar haemorrhage due to leptospirosis. Indian J Med Res. 2010;131: 429–433.

27. Dursun B, Bostan F, Artac M, Varan HI, Suleymanlar G. Severe pulmonary haemorrhage accompanying hepatorenal failure in fulminant leptospirosis. Int J Clin Pract. 2007;61: 164–167. doi:10.1111/J.1742-1241.2005.00638.X

28. Chaikajornwat J, Rattanajiajaroen P, Srisawat N, Kawkitinarong K. Leptospirosis manifested with severe pulmonary haemorrhagic syndrome successfully treated with venovenous extracorporeal membrane oxygenation. BMJ Case Rep. 2020;13. doi:10.1136/BCR-2019-230075

29. Kularathna MDSV, Kularatne SAM, Pathirage M, Nanayakkara PTMA. Severe leptospirosis complicated with multiorgan dysfunction successfully managed with plasma exchange: a case report. J Med Case Rep. 2021;15: 1–6. doi:10.1186/S13256-021-03135-3/TABLES/1

30. Kularatne SAM, Budagoda BDSS, de Alwis VKD, Wickramasinghe WMRS, Bandara JMRP, Pathirage LPMMK, et al. High efficacy of bolus methylprednisolone in severe leptospirosis: a descriptive study in Sri Lanka. Postgrad Med J. 2011;87: 13–17. doi:10.1136/pgmj.2009.092734

31. Shenoy V V., Nagar VS, Chowdhury AA, Bhalgat PS, Juvale NI. Pulmonary leptospirosis: an excellent response to bolus methylprednisolone. Postgrad Med J. 2006;82: 602. doi:10.1136/PGMJ.2005.044255

32. Smith S, Liu YH, Carter A, Kennedy BJ, Dermedgoglou A, Poulgrain SS, et al. Severe leptospirosis in tropical Australia: Optimising intensive care unit management to reduce mortality. PLoS Negl Trop Dis. 2019;13: e0007929. doi:10.1371/JOURNAL.PNTD.0007929

33. Tunjungputri RN, Gasem MH, van der Does W, Sasongko PH, Isbandrio B, Urbanus RT, et al. Platelet dysfunction contributes to bleeding complications in patients with probable leptospirosis. PLoS Negl Trop Dis. 2017;11. doi:10.1371/JOURNAL.PNTD.0005915

34. Vieira ML, de Andrade SA, Morais ZM, Vasconcellos SA, Dagli MLZ, Nascimento ALTO. Leptospira infection interferes with the prothrombinase complex assembly during experimental leptospirosis. Front Microbiol. 2017;8: 248372. doi:10.3389/FMICB.2017.00500/BIBTEX

35. Pea L, Roda L, Boussaud V, Lonjon B. Desmopressin Therapy for Massive Hemoptysis Associated with Severe Leptospirosis. https://doi.org/101164/rccm200205-450CR. 2012;167: 726–728. doi:10.1164/RCCM.200205-450CR

